# Development of Machine Learning Models to Predict Hypoglycemia and Hyperglycemia on Days of Hemodialysis in Patients with Diabetes based on Continuous Glucose Monitoring

**DOI:** 10.1101/2025.10.24.25338707

**Authors:** Mathias Kirk Lausen, Sarah Steiner Clausen, Maja Holm Bak, Inger Vestergaard Kristensen, Morten Hasselstrøm Jensen, Peter Vestergaard, Sisse Heiden Laursen, Simon Lebech Cichosz

## Abstract

**Background/Objectives:** Patients with diabetes undergoing hemodialysis (HD) are at risk of asymptomatic hypo- and hyperglycemia within 24 hours of dialysis. Continuous glucose monitoring (CGM) can improve glycemic control, and machine learning offers a promising approach to detect and predict glycemic excursions based on CGM data. This study aimed to develop machine learning models to predict substantial hypo- and hyperglycemia on dialysis days using CGM data and baseline characteristics.

**Methods:** Using data from 21 patients with diabetes receitarving HD, three classification models (Logistic Regression, XGBoost, and TabPFN) were trained and tested. Predictive features included CGM-derived metrics, HbA1c levels, and insulin use. A binary classification approach was used to predict level 2 hyperglycemia and level 1 hypoglycemia based on international consensus targets; CGM derived Time Above Range (TAR) ≥10% and Time Below Range (TBR) ≥1%.

**Results:** A total of 555 dialysis days were included in the analysis. The Logistic Regression model achieved the best performance for predicting hyperglycemia (F1 score: 0.85 [CI_95_,0.75-0.91]; ROC-AUC: 0.87 [CI_95_,0.78-0.93]). For hypoglycemia, TabPFN performed best (F1 score: 0.48 [CI_95_,0.26-0.69]; ROC-AUC: 0.88 [CI_95_,0.77-0.94]).

**Conclusion:** Prediction of substantial hypo- and hyperglycemia in patients with diabetes undergoing HD appears feasible using machine learning models. Additional studies are needed to confirm clinical utility and generalizability.

## 1. Introduction

Diabetic nephropathy occurs in 20-50% of individuals with diabetes and is the most common cause of chronic kidney disease [1–3]. Consequently, many patients progress to renal replacement therapy, with an incidence of 145 per million population in Europe in 2021 [4]. Despite advances in treatment, patients with diabetes undergoing hemodialysis (HD) have a markedly reduced survival rate of approximately 3.7 years — nearly half that of patients without diabetes on HD [5]. This underscores the critical need to optimize management strategies in this vulnerable population. Maintaining optimal glycemic control among patients on HD is therefore essential to prevent the progression of microvascular and macrovascular complications and to prolong life expectancy [6, 7]. However, managing glycemic control in patients with diabetes on HD is particularly challenging. HD itself is an independent risk factor for increased glycemic variability due to the removal of glucose and insulin during dialysis [7, 8]. Furthermore, these patients experience significant glucose fluctuations, with a higher incidence of hypoglycemia compared to patients with diabetes not receiving HD, contributing to increased morbidity and mortality [8–10]. Studies have demonstrated marked differences in glucose levels on dialysis days compared to non-dialysis days, with the highest risk of asymptomatic hypoglycemia occurring 24 hours following the beginning of dialysis and the majority of hypoglycemic episodes occurring during dialysis [8, 9, 11–14]. In addition to the intra-dialytic risk of hypoglycemia, patients also face increased risk of post-HD hyperglycemia [8, 9].

Continuous glucose monitoring (CGM) effectively detects both hypo- and hyperglycemia, providing continuous assessment of blood glucose variability [7, 15]. Use of CGM in patients with diabetes undergoing HD has been shown to improve glycemic control by reducing episodes of extreme glycemic events and decreasing glycemic variability, thereby increasing Time in Range (TIR) [7, 13–16]. CGM devices primarily provide reactive interventions by alerting users when their glucose levels fall outside target ranges. However, proactive glucose management strategies enable patients to anticipate and mitigate risk through planned interventions such as insulin administration, physical activity, and dietary adjustments [6, 17–20]. The Advanced Technologies & Treatments for Diabetes (ATTD) Congress established an international consensus on glycemic targets for people with diabetes, emphasizing increasing TIR as a primary goal. For patients with diabetes and coexisting renal disease, recommended targets include ≤1% of time below 70 mg/dL, ≤10% above 250 mg/dL, and ≥50% within the range of 70–180 mg/dL [21, 22]. Complementing this, the American Diabetes Association recommends utilizing parameters such as Time Below Range (TBR) and Time Above Range (TAR) for insulin dose adjustments and treatment regimen reevaluation, while Kidney Disease Improving Global Outcomes (KDIGO) (2022) advocates for using TIR and TBR as glycemic targets [6, 18].

Machine learning has emerged as an effective tool for risk prediction by identifying complex patterns and non-linear relationships within large datasets [20, 23]. Several studies have demonstrated its potential to predict hypo- and hyperglycemia based on CGM data [19, 20, 24]. However, to our knowledge, no study has yet developed and introduced a prediction tool that simultaneously evaluates both hypo- and hyperglycemia in patients with diabetes undergoing HD. Only one study has applied machine learning to predict the risk of intradialytic hypoglycemia in this population using CGM data [25]. By integrating predictions of both hypo- and hyperglycemia on dialysis days, such a tool could potentially facilitate timely interventions to help patients avoid glycemic excursions, thereby improving treatment strategies and enabling patients and healthcare providers with better means to proactively manage blood glucose on dialysis days. Given the complexities of glucose management in patients undergoing HD, the increased risk of complications occurring 24 hours following the beginning of dialysis, and the established international glycemic targets, this study aims to develop machine learning models to predict significant hypo- and hyperglycemia on dialysis days using CGM data and baseline patient characteristics.

## 2. Materials and Methods

### 2.1. Study Design and Ethical Considerations

In this study, prediction models were developed, trained, tested, and compared based on data collected as part of a 16-week crossover trial (ClinicalTrials.gov ID: NCT05678712) conducted in Denmark by Steno Diabetes Center North, Aalborg University Hospital, and the Department of Health Science and Technology, Aalborg University. The use of data for developing and testing prediction models was approved by The North Denmark Regional Committee on Health Research Ethics (journal number: N-20210070). The study complied with data responsibility requirements and the General Data Protection Regulation (GDPR)[26]. Informed consent was obtained from all participating patients, and the study was conducted in accordance with the ethical principles of the Declaration of Helsinki and adhered to relevant national and institutional guidelines for research ethics.

### 2.2. Setting and Study Subjects

A total of 22 adult patients (≥18 years) who participated in the 16-week crossover trial were recruited from dialysis centers in Aalborg and Hjørring, Denmark. All patients had type 1 (T1DM) or type 2 (T2DM) diabetes mellitus, were receiving chronic HD or hemodiafiltration, and were treated with insulin.

### 2.3. Data Collection and Variables

The data collected included CGM data obtained using the Dexcom G6 system, HbA1c values, and insulin dosages prior to dialysis. Predictions were made for each dialysis day, starting at the onset of the dialysis session. These data were used to train, test, and compare different machine learning models aimed at predicting episodes of hypo- and hyperglycemia (TAR and TBR, respectively). The prediction models were developed for potential use in clinical practice to support decision-making before, during, and after dialysis sessions, with the aim of enabling more individualized care and aiming to prevent episodes of substantial hypo- and hyperglycemia.

### 2.4. Modelling Framework

An overview of the modelling and evaluation process is presented in Figure 1.

**Figure 1.**
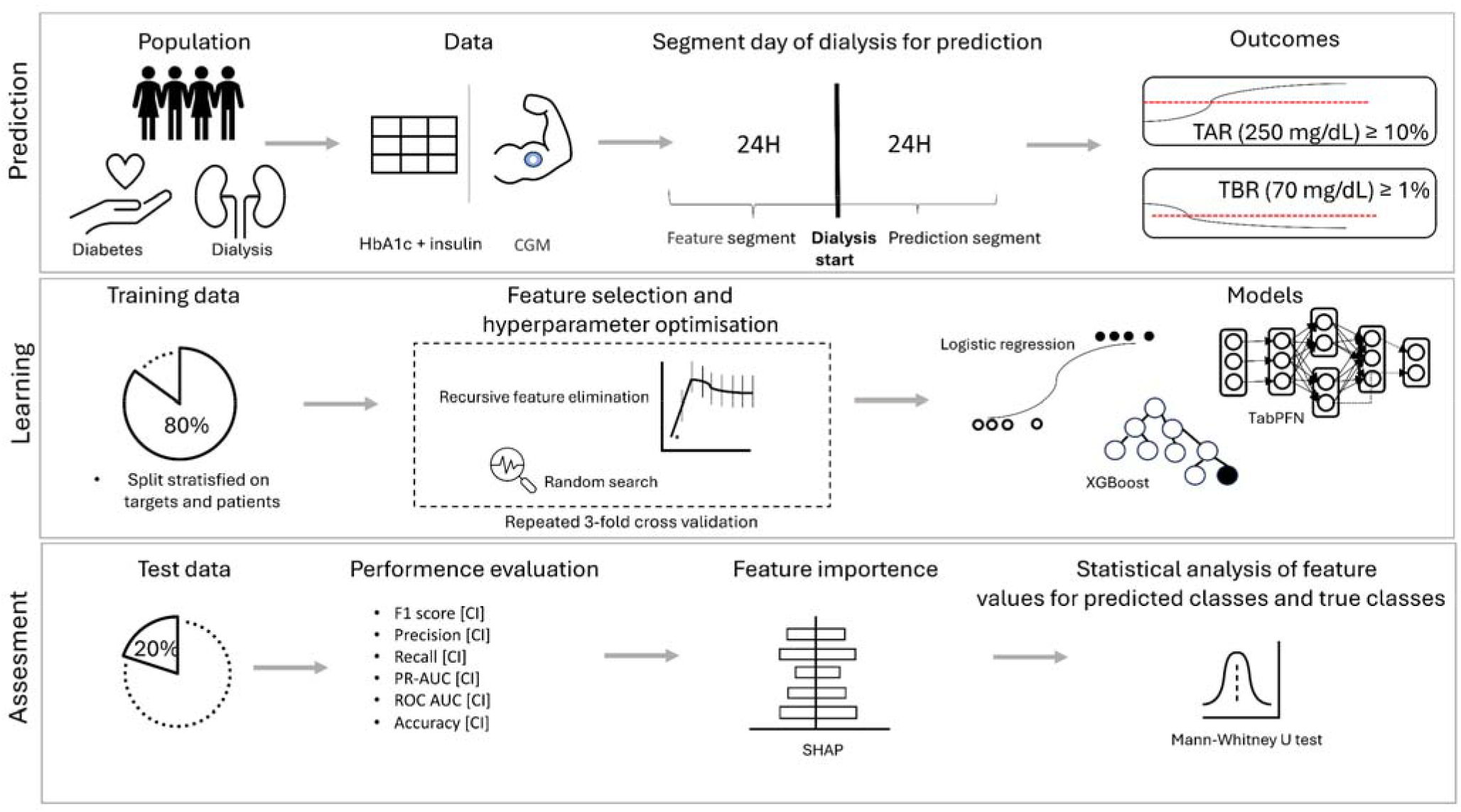
The *Prediction* row summarizes the modeling of TAR and TBR outcomes in patients with diabetes undergoing dialysis using CGM, HbA1c, and insulin data. Predictions were made for dialysis days, starting at dialysis session onset. The *Learning* and *Assessment* rows outline the model development process, including feature selection using RFECV with repeated measures and hyperparameter tuning via random search. Logistic Regression, XGBoost, and TabPFN models were subsequently trained and evaluated using performance metrics, feature importance analysis, and statistical testing.

The *Prediction* row summarizes the modelling of TAR and TBR outcomes in patients with diabetes undergoing dialysis using CGM, HbA1c, and pre-dialysis insulin dosages. Predictions were made for dialysis days, beginning at session onset.

The *Learning* and *Assessment* rows describe the modelling pipeline, including feature selection using Recursive Feature Elimination with Cross-Validation (RFECV) applied to repeated measures, and hyperparameter tuning via random search. Logistic Regression, XGBoost, and TabPFN models were trained and evaluated based on performance metrics, feature importance analyses, and statistical testing.

### 2.5. Segments

Predictions were made on the day of dialysis, starting at the onset of dialysis, targeting glycemic outcomes (i.e., hypo- and hyperglycemia) over the subsequent 24 hours. From the CGM data, segment pairs were constructed as model inputs, each consisting of a feature segment and a prediction segment. The prediction segment spanned the 24 hours following dialysis initiation, while the feature segment covered the 24 hours preceding it. Each instance was generated at the start of a dialysis session, resulting in repeated representations of individual patients, as each patient underwent multiple HD sessions. In total, data from 662 dialysis sessions were available. The segmentation approach is illustrated in the *prediction* row of Figure 1, which provides the contextual framing for the predictions.

### 2.6. Targets

The targets were defined in accordance with the ATTD (2019) recommendations for glycemic control in patients with diabetes and renal disease [21]:

- Predicted Time Above Range level 2 (pTAR2): A TAR ≥ 10% within 24 hours after dialysis initiation.
- Predicted Time Below Range level 1 (pTBR1): A TBR ≥ 1% within 24 hours after dialysis initiation.

The predictions were binary and conducted separately for the two outcomes: pTAR2 and pTBR1. Target values were derived from CGM data within the prediction segment. For each outcome, the three models were trained, tested, and compared.

### 2.7. Data Preparation and Imputation

During data collection, four patients dropped out: one before CGM data could be collected, and three after partial data collection. Data from the latter three patients were retained and included in the analysis. Thus, data from a total of 21 patients were included in the final dataset.

Across all CGM measurements, 8.5% of data points were missing, with 7.5% missing in the *feature* segment and 9.5% in the *prediction* segment. Because missing data in both segments could influence the calculated features and prediction targets, predefined inclusion and exclusion criteria were applied to all segment pairs:

1. Segment pairs were excluded if either the feature or prediction segment contained ≥30% missing CGM values [27].
2. Segment pairs were excluded if there were gaps of ≥4 consecutive hours in either segment [27].
3. Segment pairs that met the inclusion criteria (i.e., <30% missing values and no gaps ≥4 hours) were retained. Missing values in these segments were imputed using linear interpolation.

A total of 107 segment pairs were excluded based on these criteria. The remaining 555 segments pairs were linearly imputed and included in the analysis. Among these 555 instances, 317 were positive for pTAR2 and 51 for pTBR1, corresponding to class imbalance ratios of 1:1.8 and 1:10.9, respectively. For non-CGM-variables (HbA1c, long-acting insulin, and fast-acting insulin) missing values were imputed using Scikit-learn’s RandomForestRegressor (version 1.7.0) [28].

The data were randomly split into 80% training and 20% test sets using stratification based on both target variables (section 2.4.5.) and patients. Stratification at the patient level ensured that each patient appeared in only one of the two sets, while stratification on the target variables ensured balanced representation of each target class in both sets.

### 2.8. Features

A total of 37 features were derived from the CGM data within the feature segment. These features were computed using Cgmquantify (version 0.5), an open-source toolbox designed for the calculation and visualization of CGM metrics [29]. Each feature segment was divided into four consecutive subsegments of six hours each. Subsegment 1 represented the six hours immediately preceding the prediction segment, followed by subsegments 2 to 4, each covering the preceding six-hour intervals. Furthermore, patients’ pre-dialysis insulin intake and HbA1c values were included, resulting in a total of 40 features, which is shown in Table 1. The features were scaled to a range of zero to one using min-max scaling.

**Table 1.**
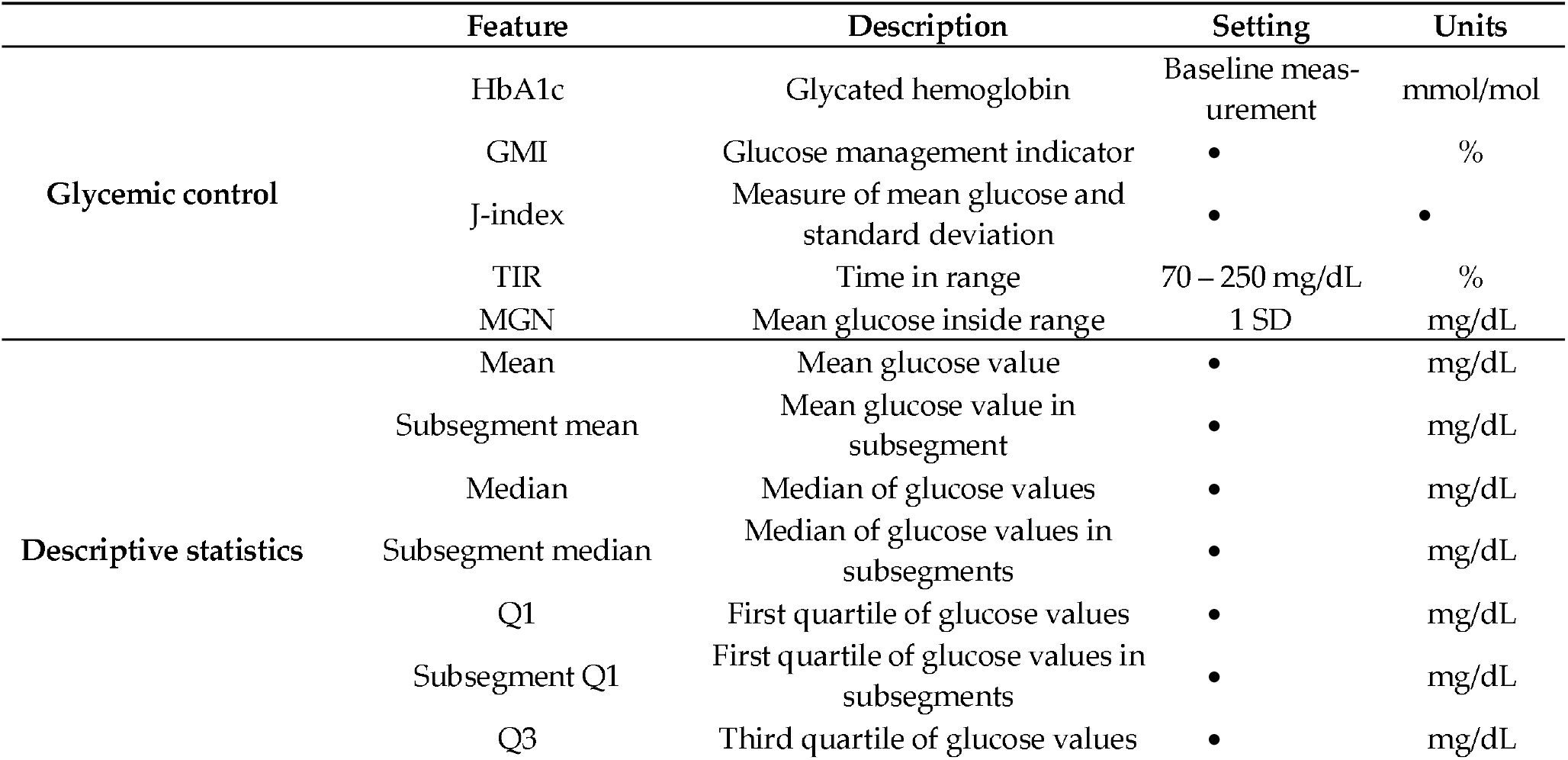
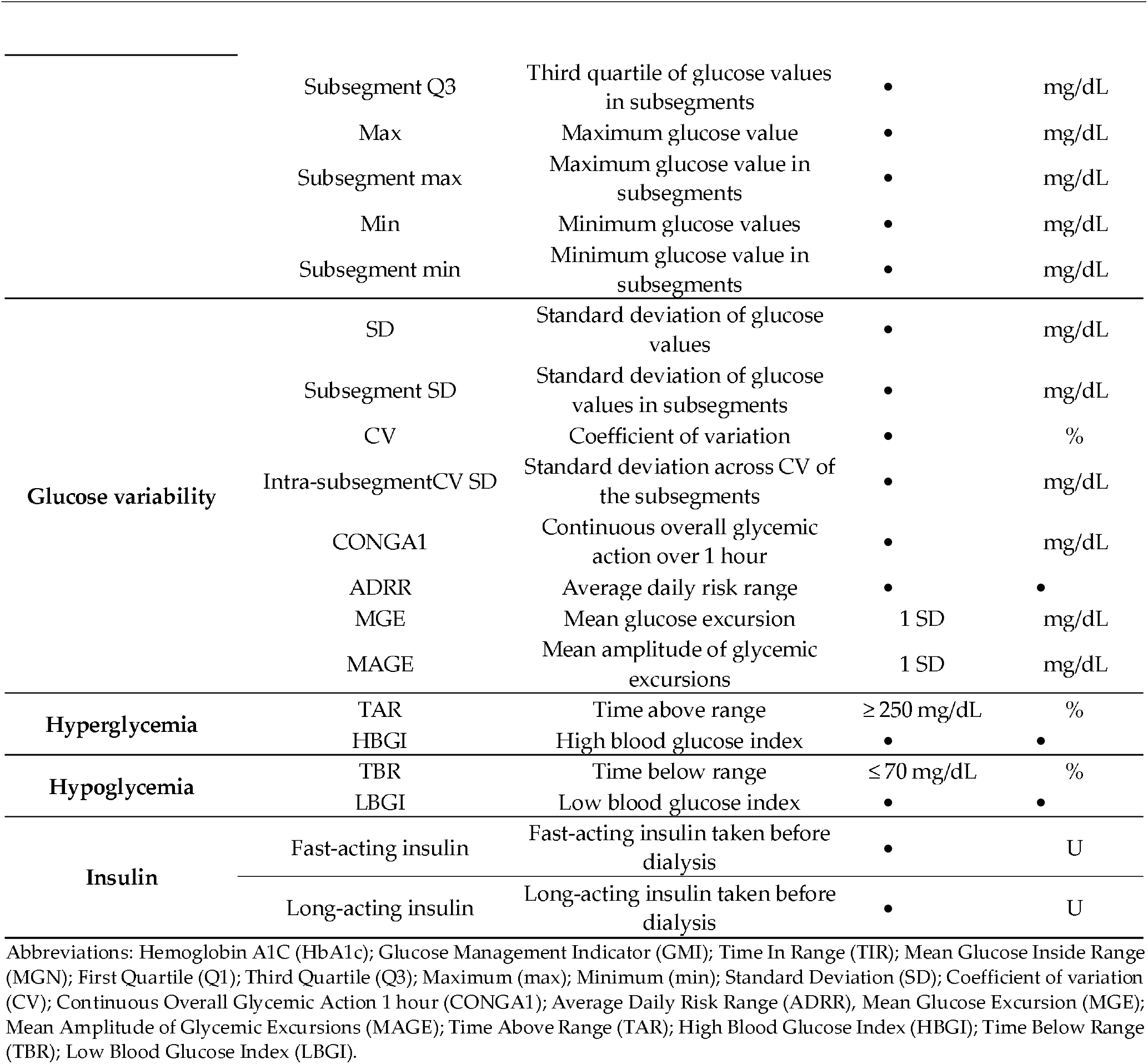
Overview of included features.

Feature selection: To select the best-performing feature subset for each model type, for both pTBR1 and pTAR2, a RFECV with five repeated measures on three-folds was used on the training data [30]. The RFECV was used with each classification model for both pTAR2 and pTBR1 to find the most optimal subset of features for each model.

### 2.9. Model Selection and Implementation Details

Three different machine learning models were evaluated for both pTAR2 and pTBR1: Logistic Regression, XGBoost, and TabPFN. These models were selected based on their distinct learning mechanisms, allowing for the identification of the most suitable approach for the prediction task.

- Logistic Regression: Logistic Regression is a generalized linear model commonly used for binary classifications [31]. It has previously demonstrated the ability to predict hypoglycemia in dialysis patients with diabetes [25, 32]. The model was implemented using LogisticRegression class from the scikit-learn package (version 1.7.0.).
- XGBoost: eXtreme Gradient Boosting (XGBoost) is a tree-based machine learning algorithm that employs boosting to improve predictive performance and is widely recognized in the field [33, 34]. XGBoost is designed to efficiently handle complex, large-scale, sparse, and imbalanced datasets, and has been shown to outperform several other machine learning algorithms [34]. The model was implemented using the XGBoost Python package (version 3.0.0.).
- TabPFN: Tabular Prior-data Fitted Network (TabPFN) is a novel and promising framework based on in-context learning, pre-trained on millions of synthetic datasets encompassing diverse prediction tasks. TabPFN has demonstrated the ability to outperform several baseline models, including XGBoost, CatBoost, and Random Forest, in classification tasks [35]. The model was implemented using the TabPFN Python package (version 2.0.8.).

The hyperparameters were optimized using random search in combination with repeated three-fold cross-validation. The cross-validation was repeated five times, and the random search was conducted over 1000 iterations. Optimization was based on the macro F1-score.

### 2.10. Model Performance Assessment

The prediction models were evaluated using the F1 score, precision, recall, PR-AUC, ROC-AUC, and accuracy for both targets: pTAR2 and pTBR1. Confidence intervals (CIs) were estimated using bootstrap resampling. The F1 score served as the primary metric for comparing model performance. Additionally, feature importance was assessed using SHapley Additive exPlanations (SHAP) [36].

### 2.11. Statistical Analysis

To analyze the distribution of feature values and assess differences between groups within the feature segment, the following comparisons were performed:

- The ground truth for TAR ≥ 10% (TAR2) group, where TAR2 comprised patients experiencing TAR2, and No-TAR2 comprised patients who did not experience TAR2 in the segment.
- The predicted pTAR2 group, where pTAR2 comprised patients predicted as positive by the model, and No-pTAR2 comprised those predicted as negative.
- The ground truth for TBR ≥ 1% (TBR1) group, where TBR1 was comprised patients experiencing TBR1, and No-TBR1 comprised patients who did not experience TBR1 in the segment.
- The predicted pTBR1 group, where pTBR1 comprised patients predicted as positive, and No-pTBR1 comprised those predicted as negative.

Statistical differences in feature values were assessed using the Mann-Whitney U test. Feature values are reported as median [Q1; Q3]. The analysis of predicted results was conducted for the best-performing model for each target. A p-value of ≤0.05 was considered statistically significant.

## 3. Results

### 3.1. Demographic and Clinical Characteristics

The demographic and clinical characteristics of the 22 study participants are summarized in Table 2.

**Table 2.**
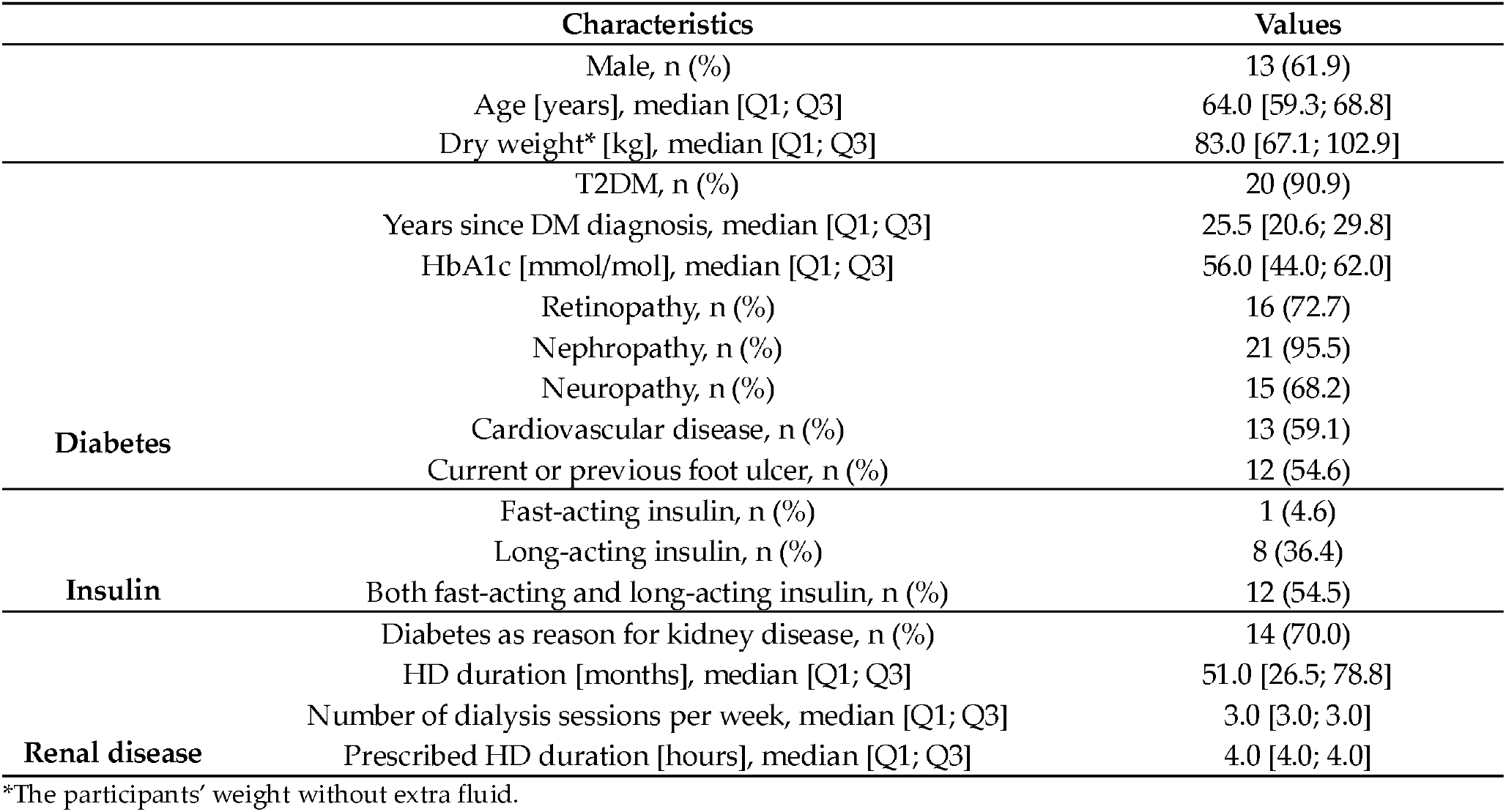
Overview of the demographic and clinical characteristics for the 22 participants at the beginning of the crossover trial.

The participants’ median [Q1; Q3] age was 64.0 [59.3; 68.8] years. 13 of the 22 participants were males. 20 participants had been diagnosed with T2DM, whereas two patients had been diagnosed with T1DM. The median [Q1; Q3] number of years since DM diagnosis was 25.5 [20.6; 29.9] years. The median [Q1; Q3] HbA1c levels were 56.0 [44.0; 62.0] mmol/mol at baseline. Diabetes complications included retinopathy, nephropathy, neuropathy, cardiovascular disease, and current or previous foot ulcers. 21 participants used an insulin pen to administer insulin and received either slow-acting insulin (n=8) or both fast-acting and slow-acting insulin (n=12). One participant received fast-acting insulin through an insulin pump, and one participant did not have their insulin prescription registered. 14 of the 22 participants had renal disease due to diabetes. The participants had undergone HD for a median [Q1; Q3] of 51.0 [26.5; 78.8] months prior to data collection, with a median [Q1; Q3] number of sessions per week of 3.0 [3.0; 3.0] and a median [Q1; Q3] of 4.0 [4.0; 4.0] hours per dialysis session.

### 3.2. Feature Selection

A total of 40 features were included prior to feature selection as shown in Table 2. Following the selection process, Logistic Regression retained 10 features for pTAR2 and 12 for pTBR1. XGBoost retained 39 features for pTAR2 and 29 for pTBR1, while TabPFN selected four features for both targets. The features included in the final models for each target are listed in Supplementary Table S1.

### 3.3. Model Performance

Table 3 presents the performance results of the three predictive models for pTAR2 and pTBR1, based on the final feature sets and test data.

**Table 3.**
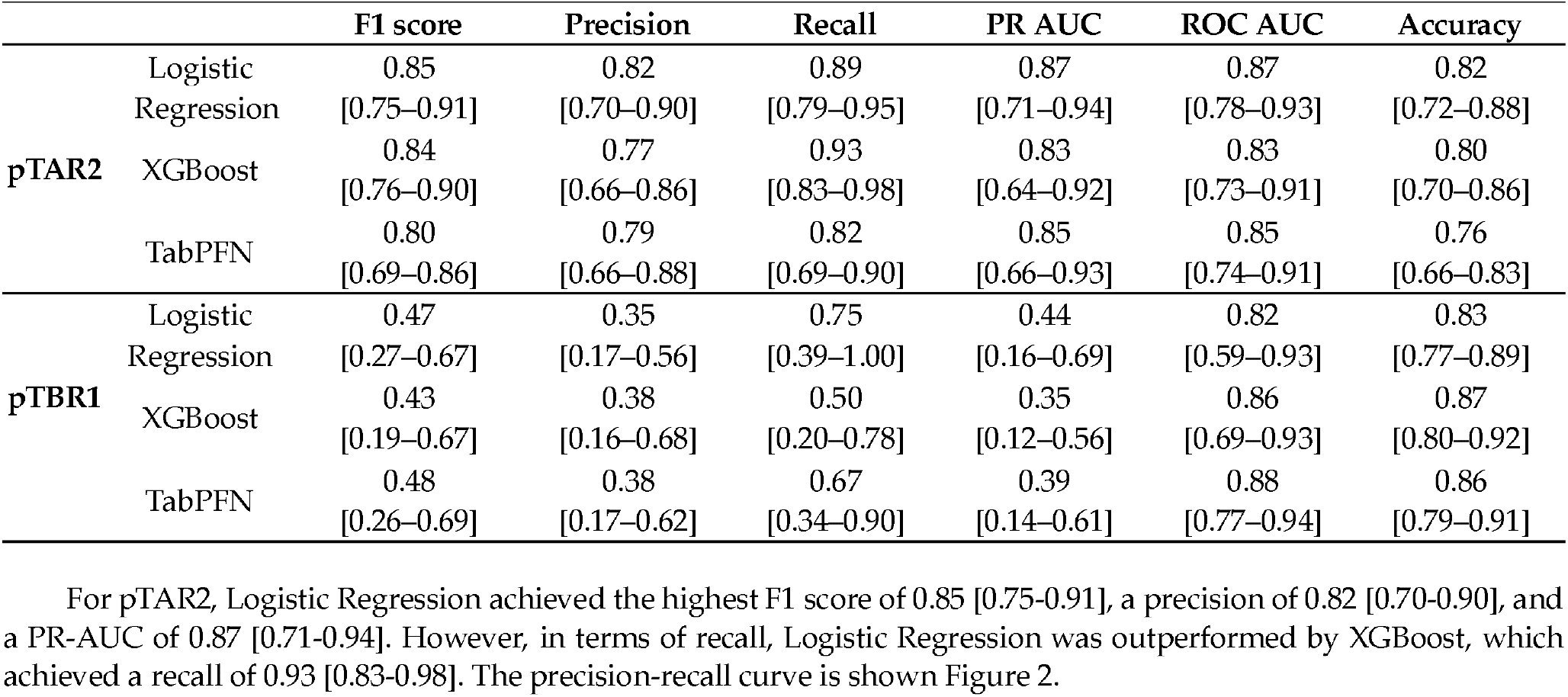
Results for the three models for targets pTAR2 (time above range level 2 ≥ 10%) and pTBR1 (time below range level 1 ≥ 1%). Results are presented with 95% confidence intervals.

Logistic Regression also achieved the highest accuracy of 0.82 [0.72-0.88] and a ROC-AUC of 0.87 [0.78-0.93]. The Receiver Operating Characteristic (ROC) curve is shown in Figure 3.

**Figure 2.**
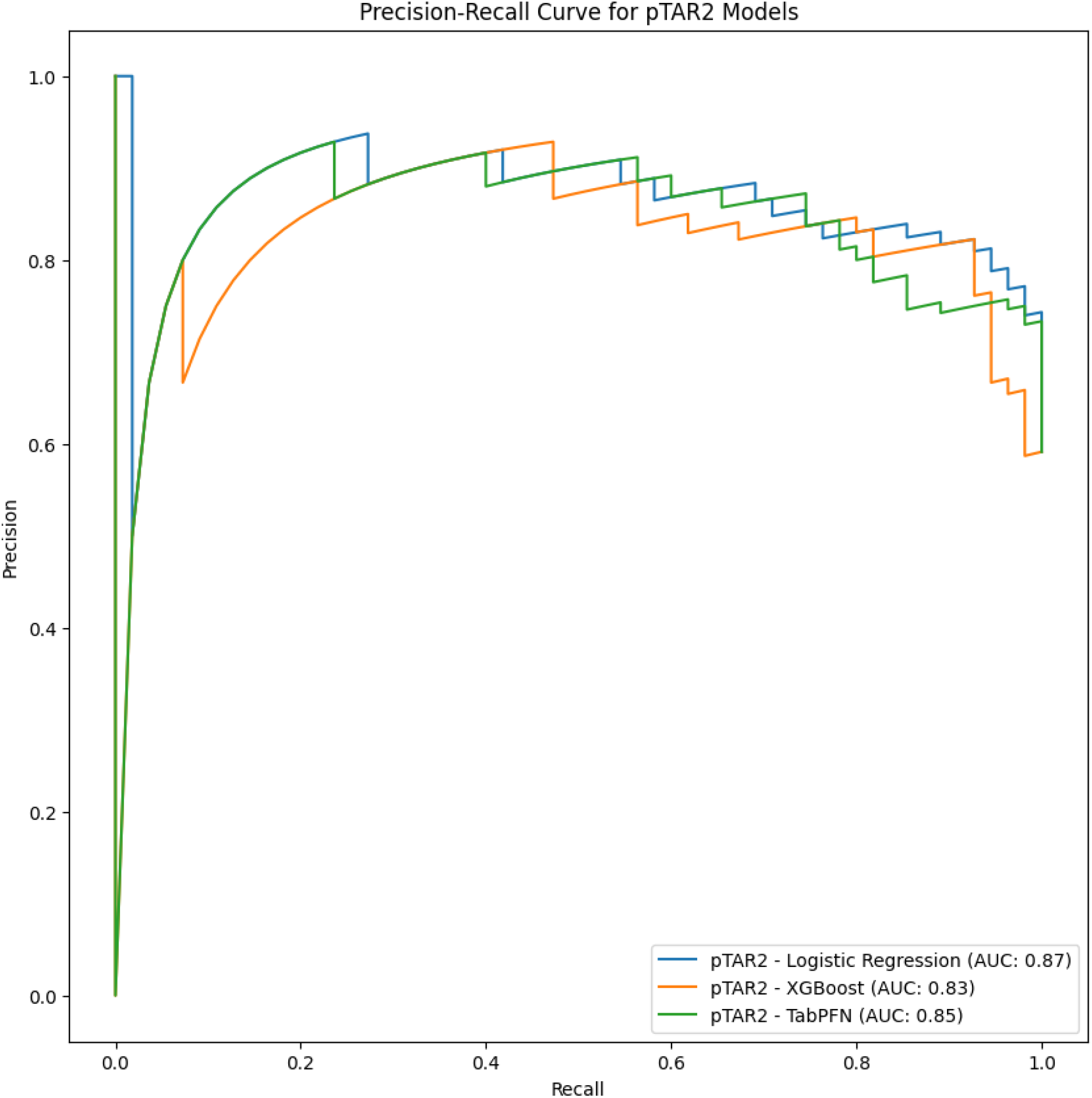
Precision-recall curves for the models predicting pTAR2 on the test set.

**Figure 3.**
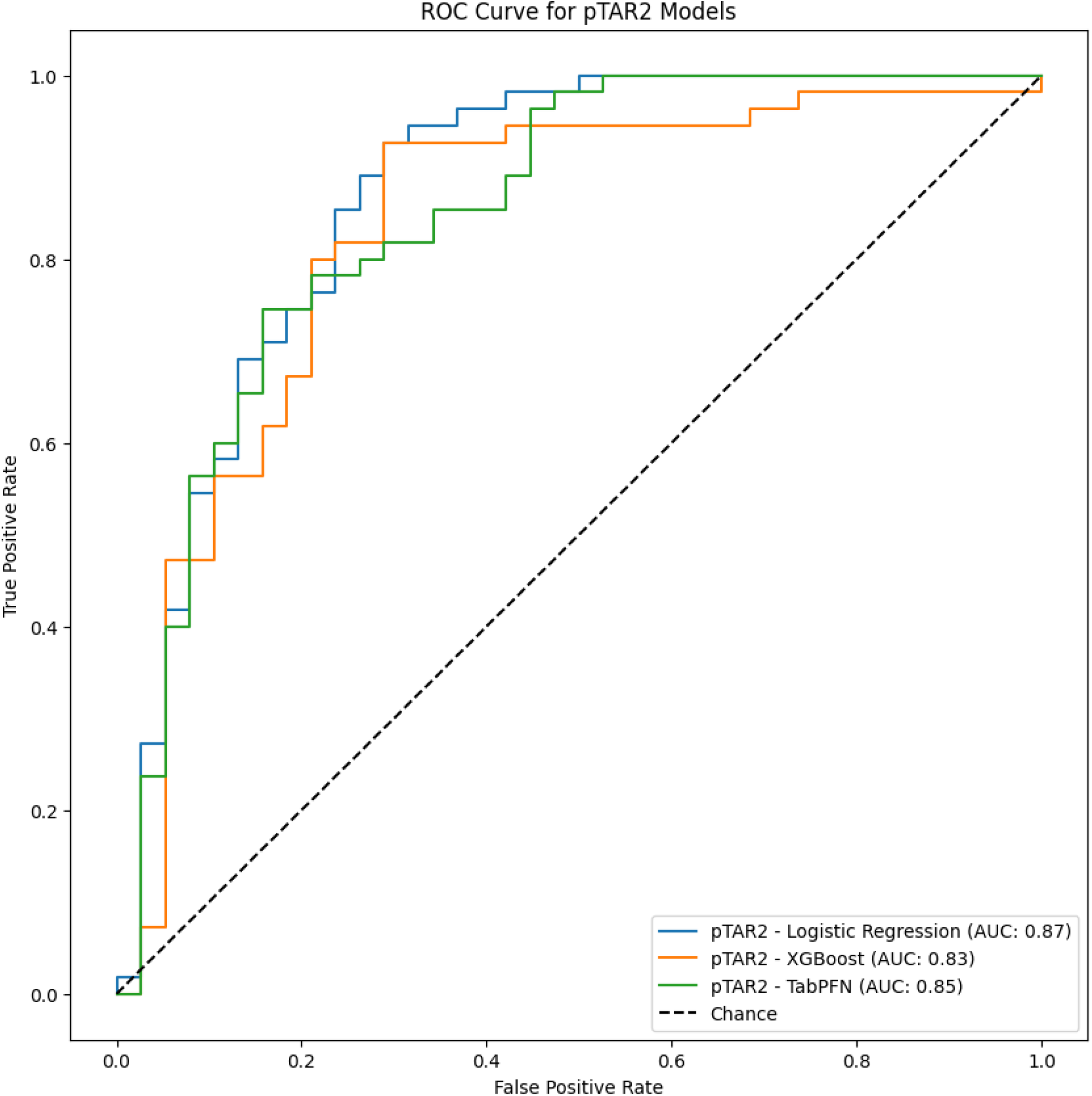
ROC curves for the models predicting pTAR2 on the test set.

For pTBR1, TabPFN was the best performing model, with an F1 score of 0.48 [0.26-0.69] and a precision of 0.38 [0.17-0.62]. In terms of recall and PR-AUC, Logistic Regression achieved the highest recall of 0.75 [0.39-1.00] and the highest PR-AUC of 0.44 [0.16-0.69]. The precision-recall curve is shown in Figure 4.

**Figure 4.**
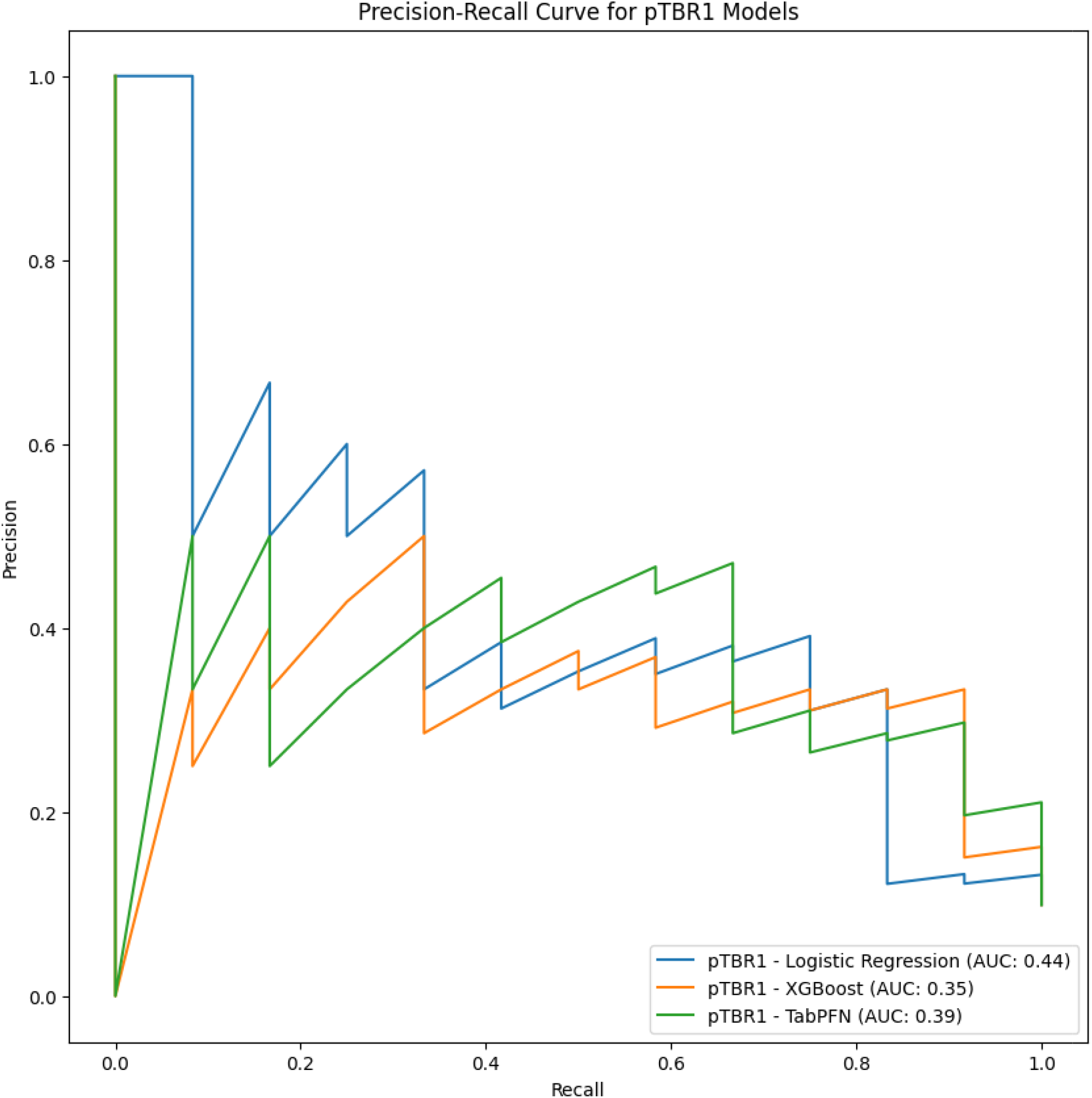
Precision-recall curves for the models predicting pTBR1 on the test set.

For pTBR1, TabPFN also achieved the highest ROC-AUC of 0.88 [0.77-0.94], while XGBoost obtained the highest accuracy of 0.87 [0.80-0.92]. The ROC curve is shown in Figure 5.

**Figure 5.**
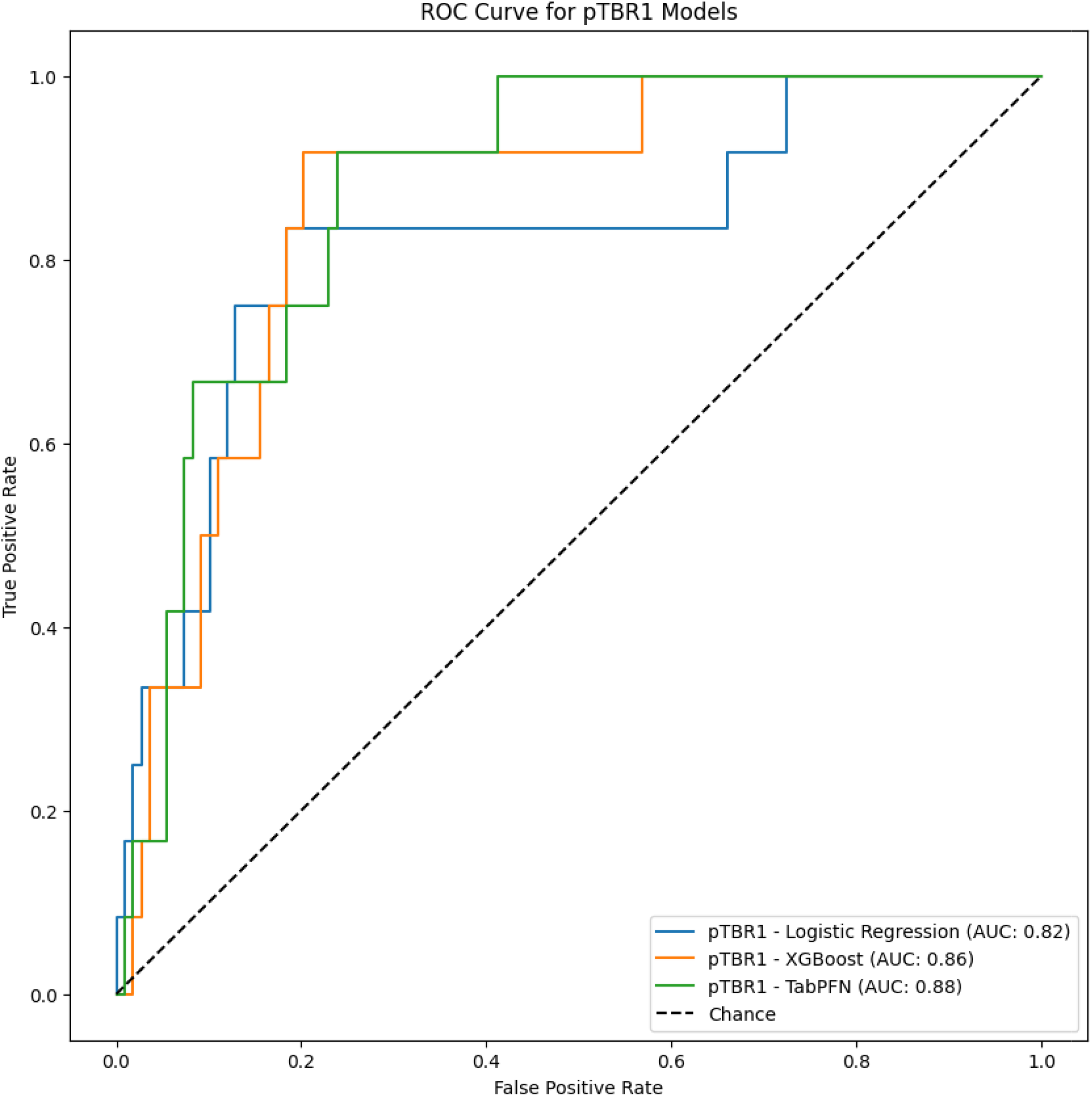
ROC curves for the models predicting pTBR1 on the test set.

Overall, the performance across all three models were relatively similar for both targets, as also reflected in the confidence intervals. The models exhibited only slight variations across the different evaluation metrics.

### 3.4. Feature Importance

Feature importance was assessed using SHAP for the best-performing models for each target. For pTAR2, Logistic Regression was the best-performing model, and the corresponding SHAP plot is shown in Figure 6. HbA1c emerged as the most important feature, with higher HbA1c values pushing the prediction toward pTAR2 and lower values pushed it away.

**Figure 6.**
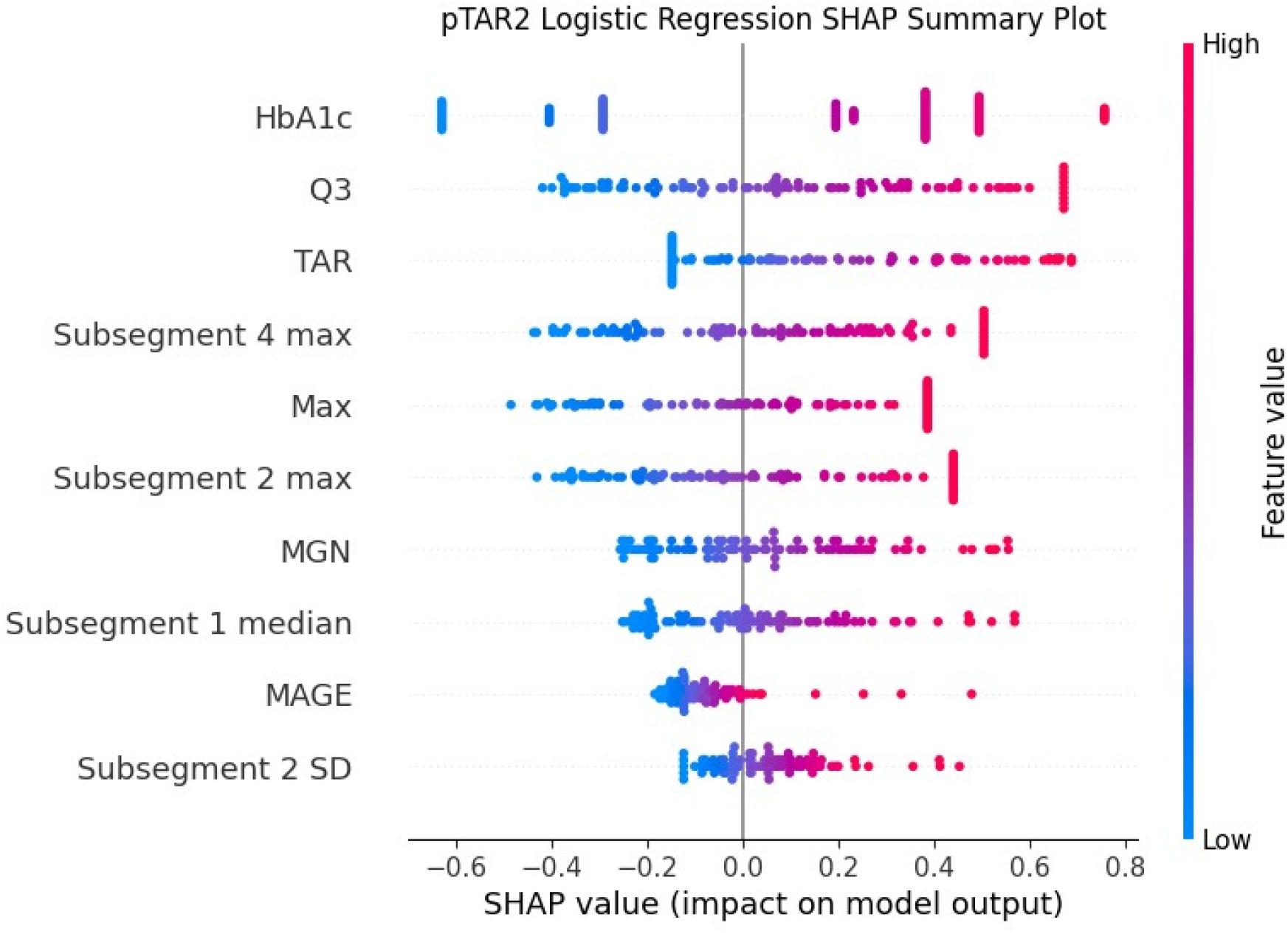
SHAP plot for the Logistic Regression model predicting pTAR2, illustrating the predictive power of each feature.

For pTBR1, the best-performing model was TabPFN; the corresponding SHAP plot is shown in Figure 7. The feature with the highest predictive power was LBGI. High values of LBGI and HbA1c pushed the prediction away from pTBR1, whereas high values of long-acting insulin and subsegment 2 median pushed the prediction toward pTBR1.

**Figure 7.**
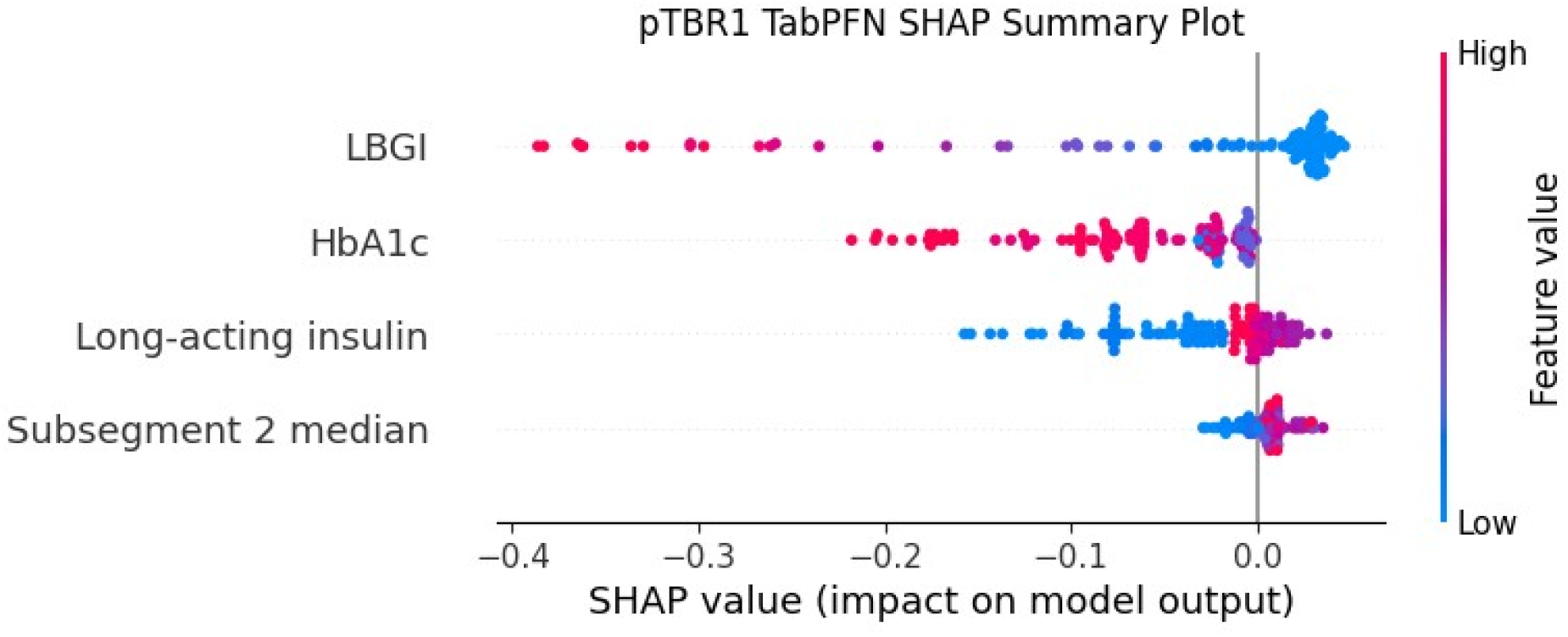
SHAP plot for the TabPFN model predicting pTBR1, illustrating the predictive power of each feature.

### 3.5. Statistical Analysis Results

The analysis was performed on the best-performing models: Logistic Regression for pTAR2 and TabPFN for pTBR1. Statistical significances and feature values are presented in Supplementary Table S2. Statistically significant differences were observed in the same features when comparing TAR2 and No-TAR2 groups as well as the predicted pTAR2 groups. For all features used in the prediction of pTAR2 by Logistic Regression (marked in blue), significant differences were found when comparing TAR2 and No-TAR2 and when comparing pTAR2 and No-pTAR2. This pattern of corresponding statistical differences was not observed in the TBR1 and pTBR1 groups. For TBR1, statistical differences were present in approximately half of the feature values when comparing TBR1 and No-TBR1 groups; however, this pattern was not replicated for pTBR1, where most features showed significant differences. Among the features used in TabPFN (marked in green), the only feature exhibiting significant differences in both TBR1 and pTBR1 was LBGI. For subsegment 1 median and Long-acting insulin, significant differences were found between the pTBR1 groups but not in TBR1. HbA1c did not show significant differences between ground truth and predicted values.

## 4. Discussion

In this study, three machine learning models were trained and tested using CGM, HbA1c, and insulin data from patients with diabetes undergoing HD to predict level 1 hypoglycemia level occurring ≥ 1% of the time or level 2 hyperglycemia occurring ≥ 10% of the time on HD days. The models demonstrated a high performance in predicting pTAR2, while predictions of pTBR1 was more challenging due to fewer hypoglycemic events and class imbalance. For pTAR2, the models showed strong discriminative ability, as indicated by F1 scores ranging from 0.80 to 0.85 and PR-AUC values ranging from 0.83 to 0.87. The predictive models for pTBR1 demonstrated limited discriminative ability, as reflected by F1 scores ranging from 0.43 to 0.48 and PR-AUC values between 0.35 and 0.44. Despite this, the results suggest that prediction models incorporating real-time CGM, insulin, and HbA1c data hold promise for personalized risk forecasting in this patient population.

### 4.1. Comparison with Previous Studies

The features with the highest predictive power varied slightly depending on the target and the model applied. However, it is noteworthy that the baseline feature HbA1c consistently demonstrated the strongest predictive value across most models. This aligns with KDIGO (2022) recommendations, which endorse HbA1c as a marker for glycemic control and maintain it as the standard of care, despite its reduced diagnostic accuracy and reliability in patients with end-stage kidney disease undergoing HD [6, 37]. In our study, HbA1c levels were, on average, lower (41.0 mmol/mol) in segments without substantial hyperglycemia and higher (62.0 mmol/mol) in segments with substantial hyperglycemia. Interestingly, segments with substantial hypoglycemia also exhibited elevated HbA1c levels (60.0 mmol/mol), suggesting that patients with higher HbA1c may be at increased risk for both hypo- and hyperglycemia. Consistent with our findings, Divani et al. (2021) [37] also reported high accuracy of HbA1c in predicting TAR. However, Divani et al. (2021) [37] did not observe satisfactory performance of HbA1c as a marker for TBR, which aligns with our findings, as no significant differences in HbA1c values were observed between segments with and without substantial hypoglycemia.

To our knowledge, this is the first study to apply machine learning to predict the risk of substantial hypo- and hyperglycemia in patients with diabetes undergoing HD on dialysis days. Piersanti et al. (2025) [25] previously predicted hypoglycemia during HD using CGM data from patients both with and without diabetes. Our study builds upon these findings by demonstrating that it is possible to predict both hypo- and hyperglycemia throughout the entire dialysis day, not solely during dialysis. In the study by Divani et al. (2021)[37], the average TAR (>250 mg/dL) over 24 hours on a HD day was reported as 9.1%, with substantial variability among participants ranging from 0.0% to 52.4%. In contrast, our data showed a higher median TAR of 13.2%, with a notably wider range extending from 0.0% to 97.2%. Regarding TBR (< 70.0 mg/dL), Divani et al. reported an average of 5.6%, with participant values ranging from 0.0% to 25.5%. In our study, the median TBR was lower (0.0%), though with a comparable range (0.0% to 22.5%). These discrepancies may reflect differences in patient characteristics, insulin management strategies, or dialysis protocols.

This study additionally compared the performance of three distinct models, revealing no substantial differences in their ability to predict the same outcome, whether pTAR2 or pTBR1. Although participant-independent splits were employed to assess model generalizability, the limited sample size of 21 participants likely reduced the statistical power to detect performance differences between models. A study by Cichosz et al. (2024) [32] employed three comparable models to predict hyperglycemia (TAR ≥ 5.0%) and hypoglycemia (TBR ≥ 4.0%) in individuals with T1DM. Their findings indicated that non-linear models, such as XGBoost and TabNet, outperformed linear models in terms of predictive performance. In contrast, our results showed no significant difference in performance between non-linear and linear models.

### 4.2. Clinical Relevance

Predictive models, such as those developed in the present study, offer a proactive and personalized approach to improving TIR, thereby potentially reducing the risk of diabetes-related complications in this population. When implemented into a clinical setting, these tools may support individualized adjustments to insulin regimens, dietary strategies, or dialysis protocols before adverse events occur, ultimately enhancing glycemic control and long-term outcomes. This proactive approach requires active engagement from both patients, who must monitor insulin use and implement treatment adjustments, and healthcare professionals, who are responsible for interpreting system outputs and providing feedback to patients. In a study by Laursen et al. (2025) [38], conducted on the same population as included in the present study, collaboration between patients and healthcare professionals was shown to enhance the benefits of CGM. A potential application of the predictive models could involve their integration into existing CGM dashboards, thereby granting patients as well as dialysis and diabetes nurses access to the model-generated predictions. Such integration could offer healthcare professionals a structured snapshot of patient-specific risk, thereby supporting targeted follow-up and facilitating timely, individualized care. However, successful implementation of such a system in clinical practice requires routine access to CGM devices for individuals with diabetes undergoing HD. Currently, CGM use is not universally established in this population, representing a key barrier to the clinical application of predictive models in this context. Previous studies have demonstrated that the use of CGM in individuals with diabetes undergoing HD can improve glycemic control [7, 13–16]. Consequently, providing access to CGM devices in combination with predictive models may further support glycemic optimization and help prevent adverse events in this population. Moreover, the capacity to predict both hypo- and hyperglycemic risk must be carefully considered to ensure that interventions aimed at mitigating one risk do not inadvertently exacerbate the other. Thus, model-generated predictions should always be interpreted alongside other relevant clinical data, and treatment decisions should account for both diabetes and renal disease.

### 4.3. Strengths and Limitations

A key strength of this study lies in its novelty; to our knowledge, it is the first to assess the risk of both hypo- and hyperglycemia using CGM data, insulin use, and HbA1c in a cohort consisting exclusively of individuals with diabetes undergoing HD, with a specific focus on the entire dialysis day. Previous studies have not employed machine learning models to predict glycemic outcomes across the full duration of a dialysis day in this population. Another strength of the study is the use of multiple machine learning model frameworks based on distinct learning algorithms, combined with an emphasis on model explainability. This approach enhances the understanding of feature relevance and provides a foundation for future research and clinical translation. However, several limitations must be acknowledged. First, the generalizability of the findings is constrained by the relatively small sample size, particularly for individuals with T1DM undergoing HD, as the dataset includes only two such cases. Due to the limited sample size, the results may be more susceptible to random variation or influenced by the specific characteristics of the included individuals. Consequently, the glycemic patterns observed in this dataset, including responses to dialysis and the distribution of hypo - and hyperglycemia, may not be fully generalizable to the broader population of patients with diabetes undergoing HD. As such, the predictive performance demonstrated in this cohort may not accurately reflect outcomes in more diverse clinical settings, thereby limiting the external applicability of the models. Future validation in larger and more heterogeneous populations is essential to establish the robustness and clinical utility of the predictive models across a wider spectrum of patients.

Another notable limitation of this study is the moderate class imbalance in the data, particularly within the hypoglycemia category. One patient accounts for 29.4% of all hypoglycemic episodes, which may lead the models to learn individual-specific patterns rather than generalizable trends across the cohort. Moreover, the limited number of hypoglycemic events observed in this study contrasts with findings reported in the existing literature [8–10, 39], suggesting that the dataset may not fully capture the expected variability in glycemic outcomes in this population. This discrepancy may, in part, be attributed to the Hawthorne effect, wherein participants alter their behavior due to the awareness of being observed. The act of wearing a CGM sensor may have heightened patients’ attention to dietary habits and insulin management, thereby reducing the occurrence of hypoglycemic events. Additionally, healthcare professionals’ awareness of the ongoing monitoring may have contributed to increased vigilance and more frequent adjustments in treatment, further limiting the occurrence of hypoglycemia during the study period.

## 5. Conclusions

We developed and evaluated machine learning models to predict the risk of substantial hypo- and hyperglycemia on HD days in individuals with T1DM and T2DM. The models incorporated CGM-derived metrics from the 24 hours preceding the HD session, insulin administered prior to the session, and HbA1c levels obtained during the baseline and washout periods of the crossover study. The findings indicate that such predictive models show promise for identifying glycemic risk in this population. However, further research is necessary to establish their clinical utility and generalizability across broader and more diverse patient populations.

## Citation

Lausen, M.K.; Clausen, S.S.; Bak, M.H.; Kristensen, I.V.; Jensen, M.H.; Vestergaard, P.; Laursen, S.H.; Cichosz, S.L.

## Author Contributions

Conceptualization, MKL., SSC., MHB., SLC., SHL., IVK., MHJ., and PV.; Methodology, MKL., SSC., and MHB.; Software, MKL., SSC., and MHB.; Validation, MKL., SSC., and MHB.; Formal Analysis, MKL., SSC., and MHB.; Investigation, SHL., IVK., MHJ., and PV.; Resources, SHL., IVK., MHJ., and PV.; Data Curation, MKL., SSC., and MHB.; Writing – Original Draft Preparation, MKL., SSC., and MHB.; Writing – Review & Editing, MKL., SSC., MHB., SLC., SHL., IVK., MHJ., and PV.; Visualization, MKL., SSC., and MHB.; Supervision, SLC., and SHL.; Project Administration, MKL., SSC., and MHB.

## Institutional Review Board Statement

In this study, prediction models were developed, trained, tested, and compared based on data collected as part of a 16-week crossover trial (ClinicalTrials.gov ID: NCT05678712) conducted in Denmark by Steno Diabetes Center North, Aalborg University Hospital, and the Department of Health Science and Technology, Aalborg University. The use of data for developing and testing prediction models was approved by The North Denmark Regional Committee on Health Research Ethics (journal number: N-20210070). The study complied with data responsibility requirements and the General Data Protection Regulation (GDPR).

## Funding

This research received no external funding.

## Informed Consent Statement

Informed consent was obtained from all participating patients, and the study was conducted in accordance with the ethical principles of the Declaration of Helsinki and adhered to relevant national and institutional guidelines for research ethics.

## Data Availability

Data cannot be published because it contains sensitive and identifying patient information. Access to the patient data through a remote desktop connection can be obtained after acceptance of a research/development/quality project application by the local approval body at Aalborg University. Contact information: Forskningsdata og Statistik, Forskningens Hus, Sdr. Skovvej 15 9000 Aalborg, Denmark.

## Conflicts of Interest

The authors declare no conflicts of interest.

## Disclaimer/Publisher’s Note

The statements, opinions and data contained in all publications are solely those of the individual author(s) and contributor(s) and not of MDPI and/or the editor(s). MDPI and/or the editor(s) disclaim responsibility for any injury to people or property resulting from any ideas, methods, instructions or products referred to in the content.

## Notes

### Competing Interest Statement

The authors have declared no competing interest.

### Author Declarations

In this study, prediction models were developed, trained, tested, and compared based on data collected as part of a 16-week crossover trial (ClinicalTrials.gov ID: NCT05678712) conducted in Denmark by Steno Diabetes Center North, Aalborg University Hospital, and the Department of Health Science and Technology, Aalborg University. The use of data for developing and testing prediction models was approved by The North Denmark Regional Committee on Health Research Ethics (journal number: N-20210070). The study complied with data responsibility requirements and the General Data Protection Regulation (GDPR). Informed consent was obtained from all participating patients, and the study was conducted in accordance with the ethical principles of the Declaration of Helsinki and adhered to relevant national and institutional guidelines for research ethics.

